# Outcomes of Venom-Induced Consumption Coagulopathy Following Snakebite Envenoming in Sudan: A Cohort Study

**DOI:** 10.64898/2026.01.26.26344815

**Authors:** Abdallateif A. Omer, Abdelsalam M.A. Nail, Burhan A. Mohammed, Rayan Ali Tonga, Tarig Elsammani Eisa, Fatima Altahir, Rania M. H. Baleela, GadAllah Modawe

**Author notes:** **Corresponding author** Dr. Rania Mohamed Hassan Baleela, Toxic Organisms Research Centre and Department of Zoology, Faculty of Science, University of Khartoum, Sudan, Email address.

## Abstract

**Background:** Snakebite envenoming (SBE) remains a major neglected tropical disease in Sudan. Venom-induced consumption coagulopathy (VICC) is the most frequent and fatal systemic complication, particularly following envenoming by hemotoxic *Echis* species. Robust clinical data on VICC in Sudan are limited.

**Methods:** We conducted a prospective hospital-based cohort study at Sinja Teaching Hospital, Sennar state, Sudan, from March to September 2022. All patients admitted with SBE were enrolled. VICC was diagnosed using the 20-minute whole blood clotting test (WBCT20) and laboratory coagulation assays. Clinical features, laboratory abnormalities, management, and outcomes were recorded until discharge or death.

**Results:** Among 119 patients with SBE (mean age 34.5 ± 9 years; 79.8% male), VICC developed in 96 (80.7%). *Echis* spp. were implicated in 86.6% of cases based on patient recognition. Spontaneous systemic bleeding occurred in 88.5% of VICC patients, and life-threatening hemorrhage in 30.2%, most commonly intracerebral hemorrhage. Acute kidney injury occurred in 36.5% of VICC cases. WBCT20 was positive in all VICC patients and showed high diagnostic sensitivity. Despite administration of fresh frozen plasma, mortality among VICC patients was 30.2%. All paediatric patients died.

**Conclusions:** VICC was highly prevalent and associated with severe hemorrhage, acute kidney injury, and high mortality in this snakebite-endemic region of Sudan. Supportive therapy alone was insufficient to prevent fatal outcomes, reflecting delayed presentation and the absence of effective *Echis*-specific antivenom. Improved access to species-appropriate antivenom, early referral, and adherence to evidence-based management are critical to reducing snakebite-related mortality in Sudan.

**Author Summary:** Snakebite envenoming is a neglected tropical disease that disproportionately affects rural and agricultural communities in low-resource settings. In Sudan, snakebite remains a major but underreported cause of illness and death. One of its most serious complications is venom-induced consumption coagulopathy (VICC), a disturbance of blood clotting that can lead to severe bleeding and organ failure.

We studied all patients admitted with snakebite envenoming to a teaching hospital in southeastern Sudan over six months. More than 80% of patients developed VICC, most often following bites attributed to *Echis* species, which are common in this region. Many patients experienced spontaneous bleeding, and nearly one-third developed life-threatening hemorrhage, most frequently bleeding in the brain. Acute kidney injury was common. Despite supportive treatment, almost one-third of patients with VICC died, and all children in the study died.

Our findings highlight the severe and largely preventable burden of snakebite envenoming in this setting. Delayed presentation to hospital, reliance on traditional healers, and the lack of effective antivenom against locally prevalent snake species contributed to poor outcomes. This study highlights the urgent need to improve access to appropriate antivenom, strengthen health-care systems, and implement evidence-based management of snakebite envenoming to reduce avoidable deaths and disability in Sudan.

## 1. Introduction

Snakebite envenomation (SBE) is one of the most significant Neglected Tropical Diseases (NTDs) in terms of incidence, severity, and clinical impact. It was recognized by the World Health Organization as one of the Neglected Tropical Diseases in 2017 [1, 2]. This was followed by the adoption of World Health Assembly resolution WHA 71.5 in 2018 and the subsequent launch of a global strategy in 2019 [3, 4] aimed at halving the burden of snakebite envenoming by 2030 [5].

In Sudan, 63,160 cases of snakebite were reported between 2014 and 2018, with an average annual incidence of 12,632 cases. The reported mortality rate among hospitalized patients during this period was 2.5% [6]. However, these figures likely underestimate the true burden of snakebite due to incomplete reporting and inadequate surveillance systems.

Most snakebite victims in Sudan are agricultural workers and children, and the incidence increases during and following the rainy season, when snake activity is higher [6, 7]. Accurate estimation of snakebite incidence remains challenging, as many individuals seek care from traditional healers rather than medical facilities. Additionally, some victims die shortly after envenoming and are therefore not captured in hospital-based records, further contributing to underreporting.

Snakebite envenoming can cause both local and systemic consequences [8]. Local manifestations include skin damage such as blisters and necrosis, cellulitis, abscesses, and myonecrosis, which may occasionally require amputation [9]. Systemic complications include coagulopathy, nephrotoxicity, cardiotoxicity, myotoxicity, and neurotoxicity [10]. Among these, coagulopathy is the leading cause of death following snakebite and a major contributor to overall morbidity and mortality= [11]. Venom-induced consumption coagulopathy (VICC) is the most common procoagulant disorder following a SBE. It results from the activation of the clotting cascade at multiple factors points specific to the toxin of the offending snake species. The primary and potentially fatal consequence of VICC is hemorrhage [12]. During this process, clotting factors are consumed leaving few or none circulation, hence the term consumption coagulopathy [13, 14]. When appropriately managed, VICC develops rapidly, typically within hours of envenoming, and resolves within 24–48 hours [15]. Clinical manifestations of VICC include spontaneous bleeding such as hematuria, gum bleeding, ecchymosis, and life-threatening hemorrhages including intracerebral hemorrhage, hematemesis, and pulmonary hemorrhage [16, 17].

## 2. Methods

### 2.1 Study design and setting

This was an observational, hospital-based cohort study conducted at Sinja Teaching Hospital, Sinja City, Sennar State, Sudan. The study aimed to determine the incidence and outcomes of venom-induced consumption coagulopathy among patients with snakebite envenoming.

### 2.2 Ethical considerations

Ethical approval was obtained from the Ethical Committees of the Sudan Medical Specialization Board, The Sennar State Ministry of Health and Sinja Teaching Hospital. Written informed consent was obtained from all participants prior to enrollment.

### 2.3 Study population

All patients admitted to the medical wards with a history of snakebite between 15 March 2022 and 15 September 2022 were eligible for inclusion. A total coverage sampling strategy was used due to the limited number of cases. Patients with known pre-existing coagulopathies or those receiving anticoagulant therapy were excluded.

### 2.4 Data collection

Data were collected using a structured questionnaire that included demographic characteristics, pictures of snake species, occupation, clinical history, site of envenoming, clinical features of coagulopathy, laboratory findings, treatment received, and outcomes. All patients underwent a detailed clinical examination and relevant laboratory investigations.

### 2.5 Snake species identification

Identification of the offending snake species was attempted using pictorial aids depicting common hematotoxic snakes, provided by the herpetology curator, Sudan Natural History Museum. Species identification was based solely on patient self-report using these images, as snake specimens were not available for morphological confirmation.

### 2.6 Assessment of coagulopathy

Patients were categorized into coagulopathy and non-coagulopathy groups based on clinical manifestations and hematological parameters. Venom-induced consumption coagulopathy was assessed using the bedside 20-minute Whole Blood Clotting Test (WBCT20) and standard coagulation tests performed in the hospital laboratory.

### 2.7 Laboratory procedures

A total of 5 mL of venous blood was collected from each patient. Three milliliters were used for routine coagulation studies, and 2 mL were used for WBCT20. For WBCT20, freshly drawn venous blood was placed into a clean, dry syringe and left undisturbed at room temperature. After 20 minutes, the syringe was gently tilted once without agitation. Failure of the blood to clot, indicated by free flow of blood, was considered a positive test result.

### 2.8 Statistical analysis

All data were entered and analyzed using the Statistical Package for the Social Sciences (SPSS), version 25 (IBM Corp., Armonk, NY, USA). Categorical variables are presented as percentages.

## 3. Results

### 3.1 Patient characteristics and presentation

A total of 119 patients with SBE were included in the study. The mean age was 34.5±9 years. Most patients were male (95/119, 79.8%). Farmers constituted the largest occupational group (52/119, 43.7%), followed by herdsmen (41/119, 34.5%). Ten of the patients were paediatric cases (10/119, 8.4%) and one of them was one year old. The lower limbs were the most common site of bite (105/119, 88.2%).

Most patents presented during the rainy season (June- August) (91/119, 76.5%), while fewer cases occurred during the summer months (March- May) (28/119, 23.5%). The majority of patients (90/119, 75.6%) arrived at the hospital on the same day as the snakebite. Delayed presentation was observed in 21 patients (17.6%), primarily due to initial consultation with traditional healers. Key demographic, clinical, and outcome characteristics are summarized in Table 1.

**Table 1.** Demographic characteristics, clinical features, and outcomes of snakebite patients.

| Variable | All Patients (n=119) | VICC (n=96) | Non-VICC (n=23) |
| --- | --- | --- | --- |
| Male sex, n (%) | 95 (79.8) | 78 (81.3) | 17 (73.9) |
| Age, years (mean ± SD) | 34.5 ± 9 | 35 ± 8 | 33 ± 10 |
| Farmers, n (%) | 52 (43.7) | 46 (47.9) | 6 (26.1) |
| Herdsmen, n (%) | 41 (34.5%) |  |  |
| Paediatric cases, n (%)* | 10 (8.4%) | 10 (10.4%) | - |
| Lower limb bites, n (%) | 105 (88.2) | 86 (89.6) | 19 (82.6) |
| Same-day presentation, n (%) | 90 (75.6) | 71 (74.0) | 19 (82.6) |
| *Echis* spp. bites, n (%) | 103 (86.6) | 89 (92.7) | 14 (60.9) |
| Spontaneous systemic bleeding, n (%) | 85 (71.4) | 85 (88.5) | 0 (0) |
| Bleeding from old healed wound, n (%) | 5 (5.2) | 5 (5.2) | 0 (0) |
| Positive WBCT20, n (%) | 96 (80.7) | 96 (100) | 0 (0) |
| INR ≥1.5, n (%) | 96 (80.7) | 96 (100) | 0 (0) |
| Acute kidney injury, n (%) | 35 (29.4) | 35 (36.5) | 0 (0) |
| Response to FFP, n (%) | - | 67/96 (70)<sup>¶</sup> | – |
| Mortality, n (%) | 29/119 (24.4) | 29/96 (30.2) | 0 (0) |
| Median hospital stay (days) | 5-7 | 5-7 | 3-5 |

\* All paediatric patients died. The youngest was one year old. <sup>¶</sup> No post-discharge follow-up was conducted; therefore, the long-term renal outcomes of patients with acute kidney injury (AKI), including progression to chronic kidney disease (CKD), could not be assessed.

### 3.2 Snake identification and clinical outcomes

Snake species identification was based on patient recognition using photographic aids, without morphological confirmation. Based on this approach, 103 patients (86.6%) identified *Echis spp.* as the offending snake, while the species could not be identified in 16 cases (13.4%).

Venom-induced consumption coagulopathy developed in 96 patients, yielding an incidence of 80.7%. All patients with VICC developed local swelling at the bite site within 30–60 minutes following envenoming. Prolonged bleeding from fang marks occurred in 79 of 96 patients (82.2%) (Fig. 1), while bleeding from a previously healed wound was observed in five patients (5.2%).

**Figure 1.**
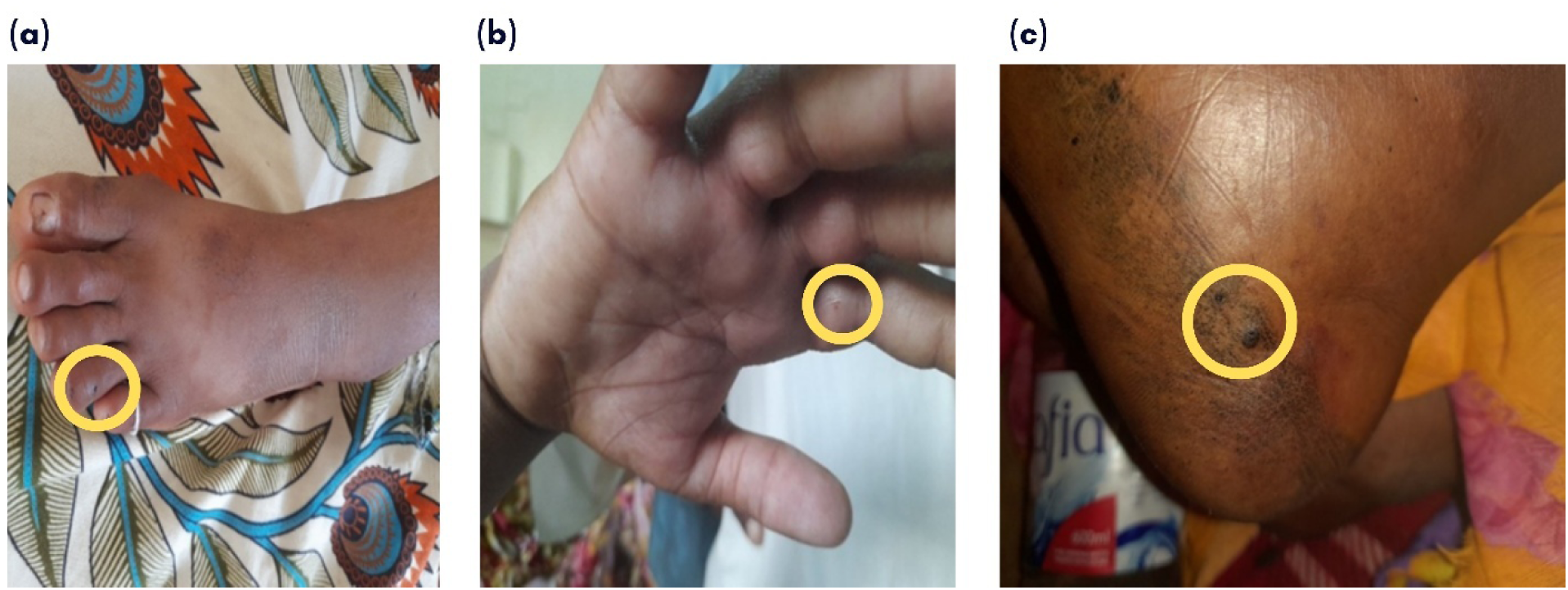
Fang marks consistent with *Echis spp.* envenoming, indicated by yellow circles: (a) left foot, fourth toe; (b) right hand, index finger; and (c) right foot, below the medial malleolus

Spontaneous systemic bleeding developed in 85 patients with VICC, frequently with multiple bleeding manifestations. These included gingival bleeding (n = 85), hematuria (n = 76), hemoptysis (n = 40), ecchymosis (n = 34, Fig. 2a), epistaxis (n = 15), subconjunctival hemorrhage (n = 14), and hematoma (n = 3). Local tissue necrosis was also observed in patients bitten by *Echis* spp. (Fig. 2b).

**Figure 2.**
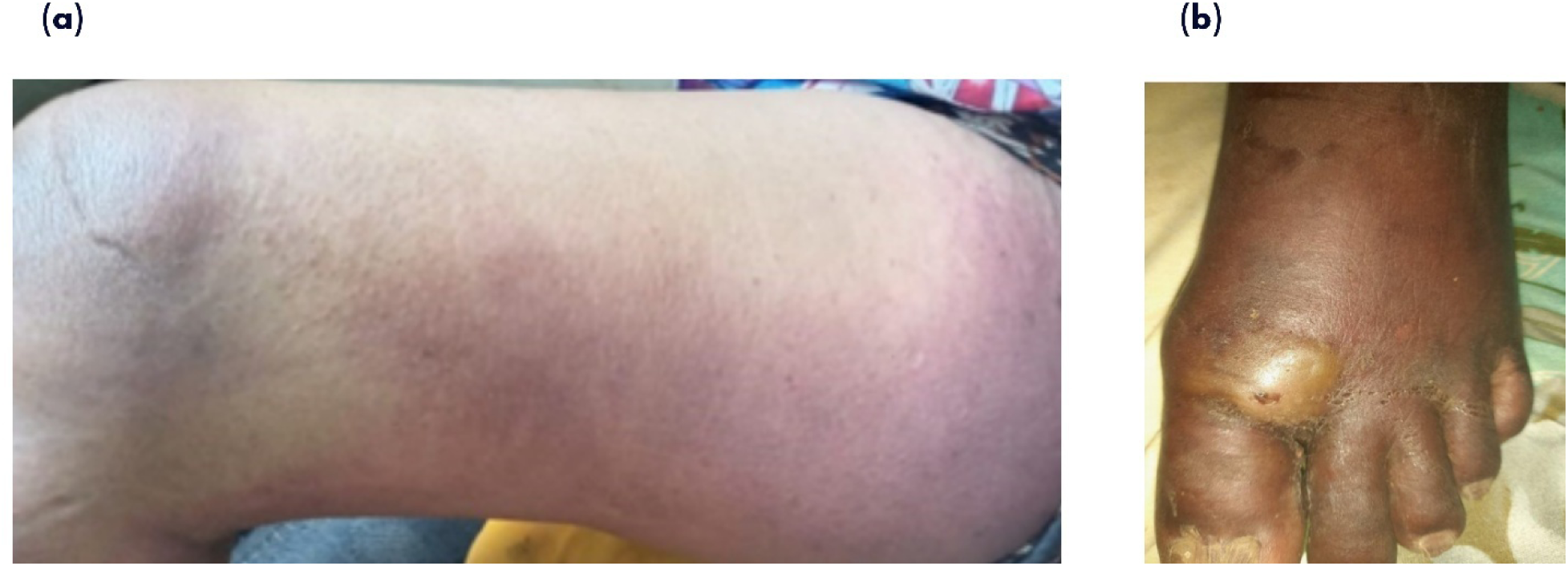
Clinical manifestations following *Echis spp.* envenoming. (a) Extensive ecchymosis over the medial thigh in a patient with venom-induced consumption coagulopathy (VICC); (b) Local tissue necrosis at the bite site on the foot following *Echis* spp. envenomation, resulting in toe amputation.

Life-threatening hemorrhage occurred in 29 patients with VICC. Intracerebral hemorrhage was the most frequent manifestation (14/29, 48.3%), followed by hematemesis (5/29, 17.2%), massive hemoptysis (4/29, 13.8%), pulmonary hemorrhage (4/29, 13.8%), and rectal bleeding (2/29, 6.9%). The onset of clinical features of VICC varied; however, most patients developed symptoms within 2–12 hours after envenoming (Fig. 3).

**Figure 3.**
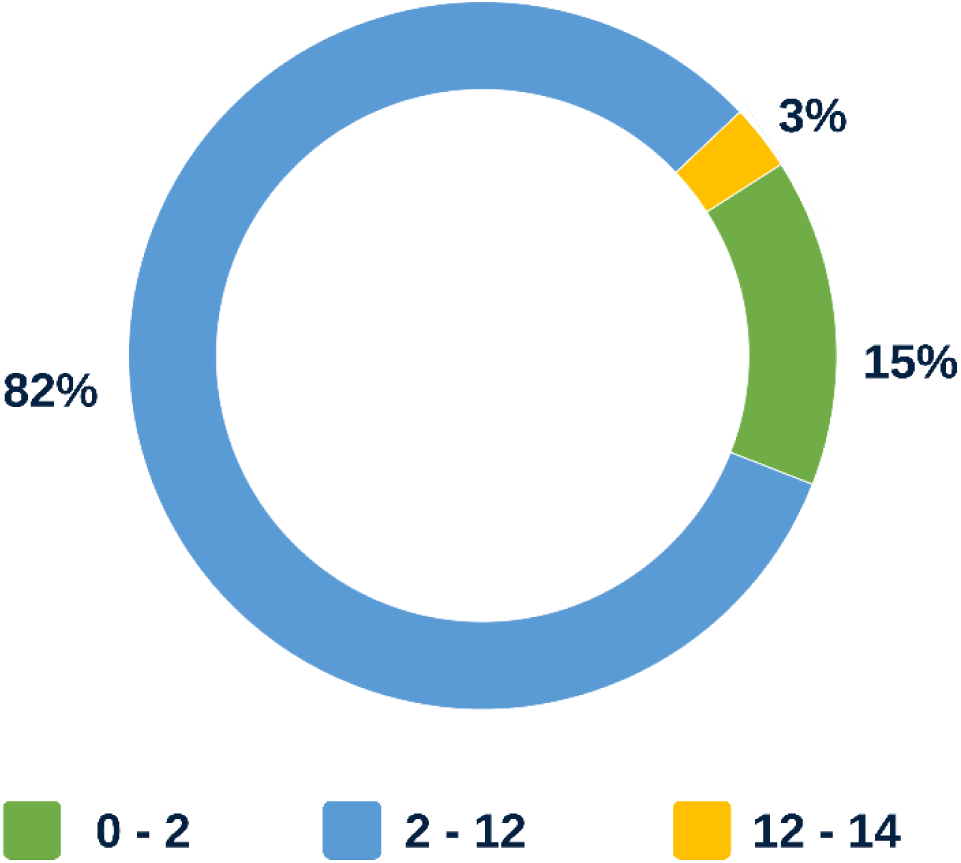
Time elapsed between snakebite envenoming and the onset of clinical features of venom-induced consumption coagulopathy (n = 96).

### 3.3 Laboratory findings and diagnosis of VICC

Venom-induced consumption coagulopathy was assessed using the bedside 20-minute whole blood clotting test (WBCT20), which was positive in all 96 patients with VICC. All VICC patients had an international normalized ratio (INR) ≥ 1.5. However, 10 patients (10.4%) demonstrated a positive WBCT20 despite an INR < 1.5, suggesting early or evolving coagulopathy.

Among patients with VICC, 35 (36.4%) had an INR > 12, 41 (42.7%) had an INR between 8 and 12, and 20 (20.8%) had an INR between 1.5 and 7. The WBCT20 showed a sensitivity of 100% and a specificity of 89.6%.

Prothrombin time was prolonged in all VICC patients (mean 117.9 seconds; maximum 270 seconds), and activated partial thromboplastin time was similarly prolonged (mean 133.5 seconds). Platelet counts remained within the normal range throughout hospitalization (mean 290 × 10³/µL). Hemoglobin levels were initially normal (mean 14 g/dL) but declined in 71 patients (74%); 12 patients (12.5%) developed hemoglobin levels < 8 g/dL and required packed red blood cell transfusion. White blood cell counts were normal in 72 patients (75%) and elevated in 24 patients (25%), with neutrophil predominance.

### 3.4 Acute kidney injury

Acute kidney injury developed in 35 of 119 patients (29.4%). According to KDIGO criteria, 15 patients were classified as stage 1, seven as stage 2, and 13 as stage 3 AKI. All patients who developed AKI had concomitant VICC.

### 3.5 Management, hospital stay, and outcomes

The antivenom supplied by the National Medical Supplies Fund (Menaven/Biosnake, Vins Bioproducts Limited [18]) is formulated to neutralize the venoms of *Naja naje, Naja nigricollis,* and *Cerastes cerastes* and does not include *Echis* spp. antivenom. In line with hospital policy, all patients with VICC received fresh frozen plasma (FFP) at 15 mL/kg, with clinical and laboratory resolution observed in 67 patients (70%). No post-discharge follow-up was conducted, so long-term renal outcomes could not be determined.

Hospital length of stay varied, with most patients discharged within 5–7 days (Fig. 4). Among patients with VICC, 67 (70%) survived and were discharged, while 29 (30.2%) died. Thirteen deaths (13.5%) occurred between days 5 and 7 of admission, five (5.2%) between days 8 and 10, and eleven (11.5%) between days 11 and 16. All paediatric cases died within 5-10 days of admission. The leading causes of death were intracerebral hemorrhage (45%) and complications related to acute kidney injury (38%) (Fig. 5).

**Figure 4.**
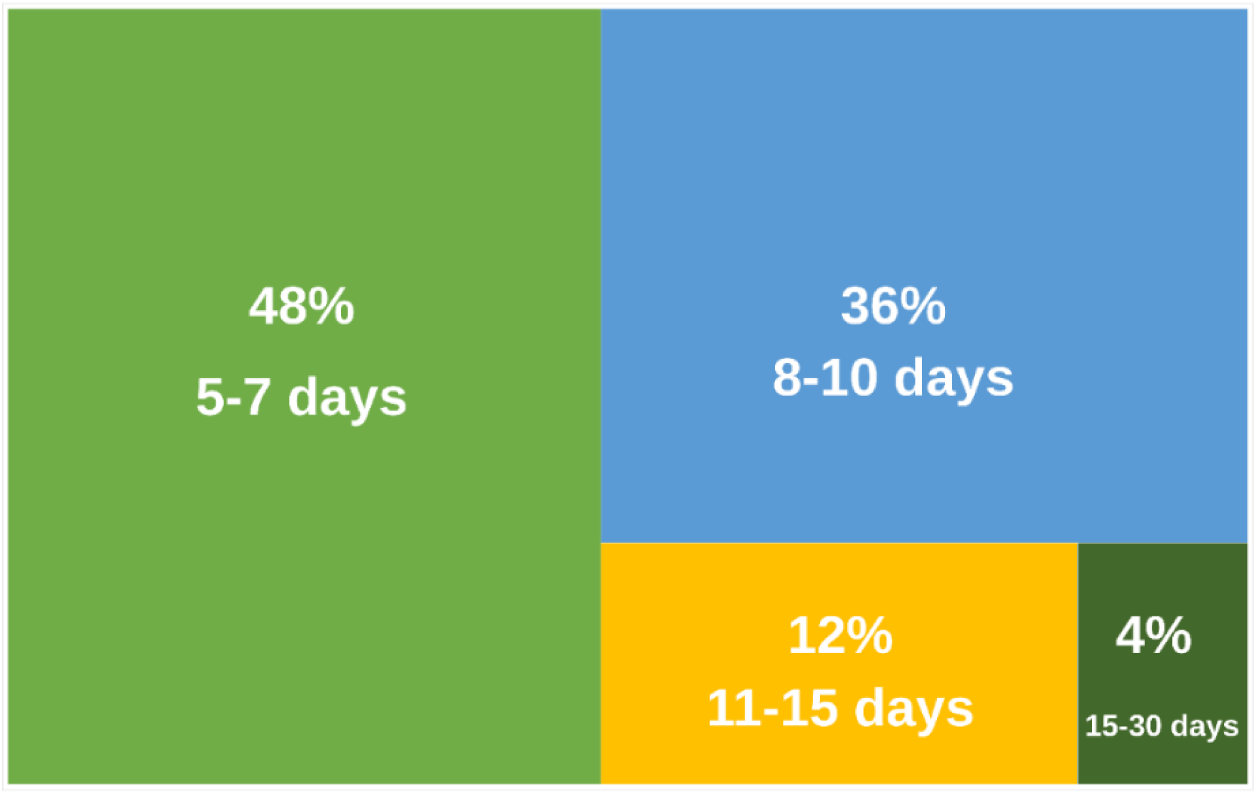
Distribution of hospital length of stay among patients with venom-induced consumption coagulopathy (VICC) (N = 67).

**Figure 5.**
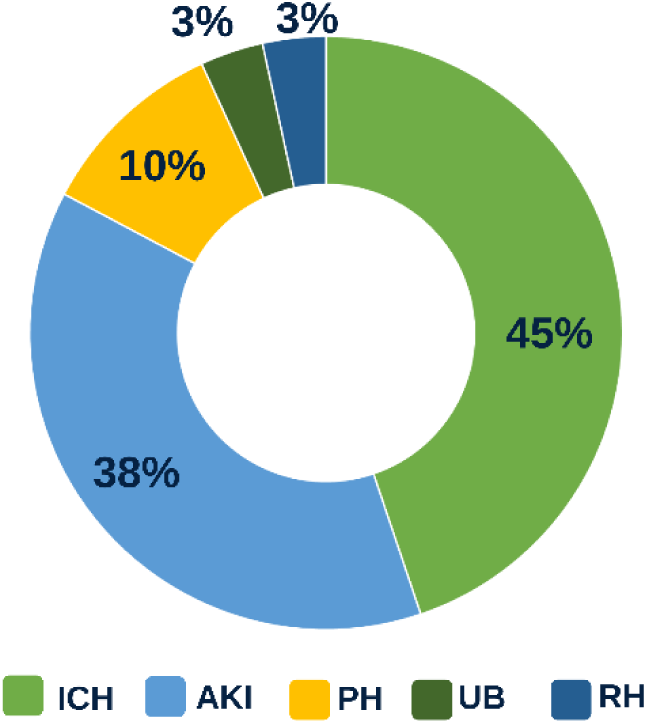
Distribution of causes of mortality among patients with VICC. Intracerebral haemorrhage (ICH) was the leading cause of death, followed by acute kidney injury (AKI), pulmonary haemorrhage (PH), upper gastrointestinal bleeding (UB), and retroperitoneal haemorrhage (RH).

## 4. Discussion

This study analysed 96 cases of venom-induced consumption coagulopathy (VICC) among 119 patients presenting with snakebite envenoming (SBE) at Sinja Teaching Hospital, located in a snakebite-endemic region of Sudan. Clinical history, physical examination, and laboratory investigations were performed on admission and monitored regularly throughout hospitalization. Patients were followed until discharge or death.

The age and gender distribution are comparable to previous reports from Sudan [6] and India [16, 17]. Most snakebites occurred among individuals aged 24 and 43, with a mean age of 34.5 ± 9 years. This reflects increased exposure among economically active adults involved in farming, livestock herding, and other outdoor activities. Seasonal variation was also consistent with studies from India [16, 17] and South Korea [19], with most bites occurring during the rainy season (June-August), when agricultural activity intensifies and snake activity increases due to emergence from shelters. In the present study, a high proportion of snakebite cases developed VICC (80.7%), exceeding rates reported in India (62% and 73%) [16, 17], South Korea (52%) [19], and Sri Lanka (33%) [20]. This elevated incidence may be attributed to the predominance of highly hemotoxic snake species in the region, delays in hospital presentation related to reliance on traditional healers and poor transportation during the rainy season, and the lack of appropriate species-specific antivenom.

A major obstacle encountered during this study was the limited availability and suboptimal species coverage of the antivenom supplied by the National Medical Supplies Fund. The available antivenom was a polyvalent formulation designed to neutralize venoms of *Naja naja*, *Naja nigricollis*, and *Cerastes cerastes*; however, it did not contain venom components from *Echis* spp., which were implicated in a substantial proportion of cases in this cohort. In addition, antivenom availability was inconsistent, with adequate supplies typically available at the beginning of each month, followed by shortages during the remainder of the month. As a result, patients were categorized into three groups based on antivenom exposure: those who received a full standard dose, those who received a partial dose, and those who did not receive antivenom.

Snake identification was performed using photographic aids depicting common hemotoxic species. Based on current distribution data, the documented *Echis* species in Sennar state is *Echis pyramidum* [21, 22].

The most frequent clinical manifestations of VICC were prolonged bleeding from fang marks, gingival bleeding, and hematuria, consistent with findings from India and South Korea [16, 17, 19]. Life-threatening hemorrhage occurred in 30% of VICC patients, which is markedly higher than rates reported in India (4–5%) and South Korea (6.2%) [16, 17, 19]. This elevated rate likely reflects delayed presentation, limited access to blood products, nursing shortages, and the absence of effective *Echis*-specific antivenom.

All paediatric patients in this cohort died, highlighting a particularly severe impact in children. Global evidence indicates higher odds of adverse outcomes in children, with a recent meta-analysis reporting a pooled case fatality rate (CFR) of 1.98% worldwide, increasing to 2.43% in sub-Saharan Africa, and 2.52-fold higher odds of adverse outcomes compared to adults [23]. Although the small number of paediatric cases limits definitive conclusions, these findings align with previous evidence that children are at increased risk of severe envenoming due to higher venom dose relative to body weight, delayed presentation, limited access to antivenom, and lack of *Echis*-specific treatment.

Laboratory abnormalities observed in VICC patients were similar to those reported in other studies [16, 17, 19]. Severe coagulopathy (INR >10) and renal impairment were associated with poor outcomes, consistent with previous evidence.

The bedside 20-minute whole blood clotting test (WBCT20) was used to diagnose VICC and was positive in all 96 cases. While all patients with positive WBCT20 had an INR ≥1.5, 10 patients had a positive WBCT20 with INR values <1.5, reflecting early or evolving coagulopathy. In this study, WBCT20 demonstrated 100% sensitivity and 98% specificity, comparable to findings from New Guinea (82% sensitivity, 98% specificity) [23] and India (94% sensitivity, 76% specificity) [24]. WBCT20 remains a practical, highly sensitive bedside test in resource-limited settings. Due to cost constraints, empty syringes were used instead of glass test tubes, which may influence clotting dynamics but reflects real-world practice.

In contrast with the available evidence and the WHO recommendations, all patients with VICC were administered fresh frozen plasma (FFP) as per the hospital regulations. Clinical and laboratory improvement were observed in 70% of VICC patients, typically within 5-7 days. In contrast, studies from India and South Korea reported lower FFP use and higher response rates, largely due to effective antivenom therapy [16, 17, 19]. The lower response rate in this study is likely attributable to the absence of *Echis*-specific antivenom. Nearly half of the patients were discharged within 5-7 days, consistent with reports from other settings.

The mortality rate among VICC patients in this study was 30%, substantially higher than rates reported in India (3–4%) and South Korea (2%) [16, 17, 19]. This high mortality rate may be partly explained by the complex pathophysiology of VICC and the limitations of current supportive treatments in Sinja Teaching Hospital, particularly the use of blood products. In the presence of un-neutralised venom procoagulant enzymes, activation of the coagulation cascade continues unabated, leading to ongoing consumption of clotting factors and, in some cases, fibrin formation. Under these circumstances, administration of blood products such as FFP may be ineffective. In resource-limited settings such as Sudan, delayed presentation, limited access to effective species-specific antivenom, and challenges in monitoring coagulation parameters may further exacerbate these risks. Together, these factors likely contributed to the persistently high mortality observed in this study.

Although 70% of patients demonstrated clinical or laboratory improvement following FFP administration, this did not translate into reduced mortality, particularly among those who developed intracerebral hemorrhage or acute kidney injury. This finding is consistent with previous studies showing that clotting factor replacement in the presence of ongoing venom activity is often transient and insufficient to prevent catastrophic bleeding.

Overall, mortality in this cohort was driven by delayed hospital presentation, envenoming by highly hemotoxic *Echis* species, absence of effective species-specific antivenom, and the development of acute kidney injury. These findings emphasize urgent needs in Sudan for improved access to appropriate antivenom, strengthened referral systems, evidence-based management guidelines, and optimized use of supportive therapies.

## 5. Limitations

Snake species identification relied on patient self-report using photographic aids, which is subject to recall bias and potential misidentification. The absence of morphological or molecular confirmation limits accurate species-level attribution. Diagnosis of venom-induced consumption coagulopathy was primarily based on the WBCT20; although appropriate for resource-limited settings, its performance may have been influenced by the use of non-standardized equipment.

Antivenom availability was inconsistent, and the available polyvalent antivenom did not cover *Echis* spp., limiting assessment of antivenom effectiveness. In addition, fresh frozen plasma was administered according to local practice despite not being recommended in the absence of effective antivenom, potentially confounding interpretation of clinical outcomes. Finally, the single-centre design and limited number of paediatric cases may restrict generalizability.

## 6. Conclusions

This study demonstrates a very high burden of venom-induced consumption coagulopathy among patients presenting with snakebite envenoming in a snakebite-endemic region of Sudan, with *Echis* spp. responsible for the majority of cases. VICC occurred in more than four-fifths of patients and was frequently complicated by severe systemic bleeding and acute kidney injury, resulting in a high case fatality rate. Mortality was predominantly driven by intracerebral hemorrhage and renal complications. Despite the use of fresh frozen plasma, clinical and laboratory improvement did not consistently translate into survival, highlighting the limited effectiveness of supportive therapy in the absence of effective species-specific antivenom. The lack of *Echis*-specific antivenom, inconsistent antivenom availability, delayed hospital presentation, and reliance on traditional healers were key contributors to poor outcomes. The uniformly fatal outcome observed among paediatric patients underscores the particular vulnerability of children to severe snakebite envenoming.

The bedside WBCT20 proved to be a highly sensitive and practical diagnostic tool for VICC in this resource-limited setting. However, effective management of snakebite envenoming in Sudan remains critically constrained by systemic gaps in antivenom access and appropriate treatment protocols. These findings emphasize the urgent need for improved availability of affordable, *Echis*-specific antivenom, strengthened referral and transport systems, early community education to reduce delays in presentation, and adherence to evidence-based management guidelines. Addressing these gaps is essential to reduce preventable morbidity and mortality from snakebite envenoming in Sudan.

## 7. Recommendations

In line with the World Health Organization (WHO) strategy to reduce snakebite envenoming–related mortality and disability by 50% by 2030, improving outcomes of venom-induced consumption coagulopathy (VICC) in this setting requires coordinated action across antivenom access, health system strengthening, community engagement, and surveillance.

Priority should be given to securing a reliable supply of quality-assured antivenom effective against locally prevalent snake species. Antivenom procurement and distribution policies should be informed by regional snake distribution and clinical epidemiology, consistent with WHO recommendations on matching antivenom to local venomous species. The absence of species-appropriate antivenom was a key contributor to poor outcomes and high mortality in this study.

Reducing delays in hospital presentation is essential to lowering morbidity and mortality. Community education and engagement programs should be strengthened to discourage reliance on traditional healers and promote early presentation to health facilities, particularly during the rainy season when snakebite incidence is highest. These efforts should be complemented by improved transportation and referral systems linking rural communities to equipped health facilities, in accordance with WHO health system strengthening objectives.

Diagnostic capacity for VICC should be reinforced through standardized use of the 20-minute whole blood clotting test (WBCT20) at peripheral and referral levels, as recommended by WHO for resource-limited settings. Where available, laboratory coagulation assays should support diagnosis and guide management. National and regional clinical management guidelines for snakebite envenoming and VICC should be developed or updated to align with WHO recommendations, including evidence-based use of antivenom, cautious use of blood products, and timely referral for severe complications such as intracranial haemorrhage and acute kidney injury.

Health facilities managing snakebite cases should be equipped with essential supplies, including antivenom and blood products, and staffed by adequately trained personnel. Regular training and continuing professional development for clinicians and nursing staff in the recognition and management of snakebite envenoming and VICC are essential components of the WHO roadmap’s capacity-building pillar.

Establishing a regional or national snakebite surveillance system is essential to monitor incidence, outcomes, and treatment effectiveness, supporting evidence-based planning and policy decisions. Future research should prioritize accurate identification of offending snake species using morphological and molecular methods to strengthen epidemiological data and guide antivenom selection, in line with WHO calls for improved data generation and research.

## List of Abbreviations

The following abbreviations and symbols were used in this manuscript:

AKI: Acute Kidney Injury
APTT: Activated Partial Thromboplastin Time
FFP: Fresh Frozen Plasma
INR: International Normalized Ratio
KDIGO criteria: Kidney Disease: Improving Global Outcomes criteria
NTDs: Neglected Tropical Diseases
PT: Prothrombin Time
VICC: Venom-Induced Consumption Coagulopathy
WBCT20: 20-min Whole Blood Clotting Test

## Declarations

### Ethics approval and consent to participate

This study was approved by the Sudan’s Medical Specialization Board, Ministry of Health Sennar state, and Sinja Teaching Hospital’s Ethical Committees.

### Consent for participation

Consent for participation was collected from all study participants or their guardians prior to their involvement in the research. Participants were informed of the study’s purpose, procedures, and potential risks. They were assured that their participation was voluntary and that their personal information would remain confidential. Written consent was obtained before data collection began.

### Consent for publication

All authors have read and agreed to the published version of the manuscript.

### Data availability statement

All data analyzed during this study are included in this published article and its supplementary information files. The raw datasets are available from the first author on reasonable request, in accordance with ethical approval and participant consent.

### Authors’ contributions

Conceptualization: **AO, AN, BM, FA, GM;** Funding acquisition: **AO;** Data curation: **AN, BM, RT, TE, FA, GM;** Investigation: **AO, AN, BM, RT, TE, FA, GM;** Methodology: **AN, BM, FA, GM;** Formal analysis: **FA, RB, GM;** Resources: **AN, BM, RT, TE;** Supervision: **AN, BM;** Visualization: **AN, BM, RB;** Writing – original draft: **AO, AN, BM, RT, TE, FA, RB, GM;** Writing – review & editing: **AO, RT, TE, FA, RB, GM**

## Acknowledgements

The authors acknowledge Ms. Entisar Khider, a herpetologist and molecular biologist at the Sudan Natural History Museum, Faculty of Science, University of Khartoum, the patients who participated in this study and the staff of Sinja Teaching Hospital, Sennar state, Sudan.

## Authors’ information

Email address, Email address, Email address, Email address, Email address, Email address, Email address, Email address

